# Medication Errors Associated with Anticoagulants at a Tertiary Care Hospital in Saudi Arabia – A Retrospective Cohort Study

**DOI:** 10.1101/2025.05.25.25328328

**Authors:** Mona Alkredees, Aseel Alsreaya, Tahani Alkredees, Khalid Ageeli, Rahaf Aljafel, Mohammed Najie, Amani Alwady, Arwa Gharsan, Bayan Alghamdi, Atheer Assiri, Rama Alsreaya, Hussam Kariri, Ali Zarea, Majid Ali

## Abstract

This study aimed to evaluate the types, frequency, severity, and contributing factors of medication errors associated with anticoagulants in a tertiary care hospital in Saudi Arabia, with particular focus on identifying patterns across different anticoagulant types and patient characteristics. A retrospective cohort study was conducted analyzing 630 anticoagulant-related medication errors reported between January 2021 and December 2023 at the Armed Forces Hospital Southern Region. Data were extracted from the hospital’s electronic reporting systems (Datix and Intervention systems). Errors were classified according to type, subtype, severity (using the NCC MERP index), and contributing factors. Statistical analyses included descriptive statistics and logistic regression to identify associations between patient characteristics and error patterns. A total of 650 medication errors were reported during the study period. Prescribing errors were the most prevalent type (72.4%), with incorrect dosing being the predominant subtype (61.3%). Enoxaparin was associated with the highest number of errors (51.2%), followed by apixaban (25.2%) and warfarin (18.2%). The majority of errors (91.4%) were classified as category B (reached the patient but caused no harm). Elderly patients (70-84 years) experienced the highest frequency of errors, with a significant association between advanced age and error severity (OR 1.42 per decade increase, 95% CI: 1.28-1.57, p<0.001). Lack of knowledge (39.7%) and monitoring failure (33.4%) were identified as the primary contributing factors, with distinct patterns observed across different anticoagulants and error types. Anticoagulant-related medication errors remain a significant patient safety concern, with prescribing errors and incorrect dosing being particularly prevalent. Targeted interventions addressing knowledge gaps, monitoring processes, and the unique challenges of anticoagulant management in elderly patients are needed to enhance medication safety and improve patient outcomes.

## Introduction

Anticoagulant medications play a vital role in modern medicine by preventing and treating thromboembolic disorders, significantly reducing morbidity and mortality associated with conditions such as atrial fibrillation, deep vein thrombosis, and pulmonary embolism. However, their narrow therapeutic index and complex dosing regimens necessitate meticulous monitoring and management, classifying them as high-alert medications [1]. Errors at any stage of anticoagulant use can lead to severe adverse events, including life-threatening bleeding or thromboembolic complications, resulting in increased morbidity, prolonged hospital stays, and potential mortality [2–4]. The economic impact of these preventable adverse drug events is substantial, underscoring the urgent need for improved safety measures [5].

The introduction of direct oral anticoagulants (DOACs) has revolutionized anticoagulation therapy by offering advantages over traditional vitamin K antagonists (VKAs) such as warfarin, including predictable pharmacokinetics, fixed dosing regimens, and reduced monitoring requirements [6,7]. Despite these advancements, medication errors persist, particularly concerning inappropriate prescribing, dose adjustments for renal impairment, and drug-drug interactions [8,9]. Prescribing errors constitute a significant proportion of anticoagulant-related medication errors, highlighting the ongoing need for improved prescribing practices and enhanced provider education [10].

Medication errors involving anticoagulants can occur throughout the medication use process. Prescribing errors often involve incorrect dosing, failure to adjust doses based on renal function, or inappropriate drug selection [11]. Dispensing and administration errors may stem from communication breakdowns, unclear labeling, or inadequate staff training. Monitoring errors, such as neglecting to track renal function or coagulation parameters, further increase the risk of adverse events, especially in patients receiving DOACs or heparins [12].

Several factors contribute to anticoagulant-related medication errors. Insufficient provider knowledge regarding appropriate dosing and monitoring requirements is a major concern. While electronic health records (EHRs) and clinical decision support systems (CDSS) aim to improve medication safety, limitations such as alert fatigue, poorly designed interfaces, or incorrect defaults can compromise their effectiveness [13]. Poor communication between healthcare providers and patients, especially during care transitions, frequently results in medication discrepancies [14]. Additionally, inadequate patient education on adherence and monitoring can lead to suboptimal outcomes.

To mitigate these risks, a multifaceted approach is necessary. Strategies such as implementing standardized protocols, enhancing provider education, and utilizing advanced technological solutions have shown promise in reducing anticoagulant-related errors [15,16]. Interventions combining education, protocol development, and technology have proven effective in addressing medication safety challenges [16]. Individualized therapy and regular assessment of renal function are particularly crucial when managing patients on DOACs, given their reliance on renal clearance.

This study aims to contribute to the growing body of knowledge on anticoagulant medication safety by analyzing medication errors associated with these drugs in a tertiary care hospital in Saudi Arabia. By examining the types of errors, contributing factors, and outcomes, we aim to provide valuable insights that can inform the development of targeted interventions to enhance patient safety and improve the quality of care for patients receiving anticoagulant therapy.

## Materials and Methods

### Study Design and Setting

This retrospective cohort study was conducted to evaluate medication errors associated with anticoagulants at the Armed Forces Hospital Southern Region (AFHSR), a tertiary care institution in Saudi Arabia. The study examined anticoagulant-related medication errors documented in the hospital’s two electronic reporting systems between January 1, 2021, and December 31, 2023. One researcher had access to the data during this period who collected the data without identifiable information of the patients.

### Study Population

The study population included all patients who had anticoagulant-related medication error reports documented in the hospital’s electronic reporting systems during the study period. The anticoagulants of interest included warfarin, direct-acting oral anticoagulants (DOACs), and heparins (both unfractionated and low molecular weight heparins).

### Inclusion and Exclusion Criteria

#### Inclusion Criteria

- Patients with documented anticoagulant-related medication errors during the study period.
- Patients prescribed any of the following anticoagulants:
- `o` Warfarin
- `o` Direct-acting oral anticoagulants (DOACs)
- `o` Heparins (unfractionated and low molecular weight heparins).
- Patients with complete electronic health records (EHR) containing the required data, including:
- Patient demographics (age, gender, diagnosis, comorbidities).
- Anticoagulant medication details (type, dose, frequency, duration).

#### Exclusion Criteria

- Patients with anticoagulant-related medication errors reported outside the study period (before 1^st^ January 2021, or after 31^st^ December 2023).
- Patients with incomplete or missing EHR data for the required variables.

### Data Collection

Data were extracted retrospectively from the hospital’s EHR and incident reporting system. The data collection form was designed to ensure standardized and comprehensive data extraction. All data were anonymized using unique study identifiers to protect patient identities, in compliance with ethical guidelines.

The following variables were collected:

- Patient demographics: Age, gender, diagnosis, and comorbidities.
- Anticoagulant medication details: Type of anticoagulant, dose, frequency, and duration of therapy.
- Medication error details: Type of error (e.g., prescribing, dispensing, administration, monitoring), severity, and contributing factors.
- Clinical outcomes: Adverse events or complications associated with the medication errors, such as bleeding or thromboembolic events.

### Outcome Measures

The primary outcomes of interest were:

1. The types and frequencies of medication errors associated with anticoagulants.
2. The severity of these errors, classified using the National Coordinating Council for Medication Error Reporting and Prevention (NCC MERP) index [17].
3. Potential contributing factors to medication errors, including patient-specific, provider-specific, and system-level factors.

### Statistical Analysis

Descriptive statistics were used to summarize the characteristics of the study population and the types and frequencies of medication errors. Subgroup analyses were performed to compare error rates and patterns between different anticoagulants (e.g., heparin, warfarin, DOACs) and by patient characteristics (age, comorbidities) and hospital units.

Univariate and multivariate logistic regression analyses were conducted to identify potential risk factors associated with medication errors. Variables included in the regression models were selected based on clinical relevance and statistical significance in univariate analyses. Adjusted odds ratios (ORs) and 95% confidence intervals (CIs) were reported.

### Ethical Considerations

Ethical approval for this study was obtained from the Institutional Review Board (IRB) of AFHSR (reference number: AFHSRMREC/2024/Pharmacy/753). Given the retrospective nature of the study, a waiver of informed consent was sought and approved by the IRB, ensuring that the research involved no more than minimal risk to subjects and could not practicably be carried out without the waiver. Patient confidentiality was maintained by anonymizing all data and storing it in password-protected files accessible only to the research team. The study adhered to the ethical principles outlined in the Declaration of Helsinki and complied with Health Insurance Portability and Accountability Act (HIPAA) regulations to safeguard patient privacy [18,19].

## Results

### Demographics and Baseline Characteristics

A total of 630 medication errors related to anticoagulants were reported during the study period, with 239 (37.9%) documented in the Datix system and 391 (62.1%) in the Intervention system. The demographic characteristics of the study population are presented in Table 1. The mean age of patients was 67.5 years (SD = 17.8, range 24-84 years), with the majority of errors reported in the age group 70- 84 years (n = 217, 34.4% of all errors), as illustrated in Fig 1. Female patients constituted 52.87% (n = 332) of the reported errors, while males accounted for 46.97% (n = 295), as shown in Table 1.

**Fig 1.**
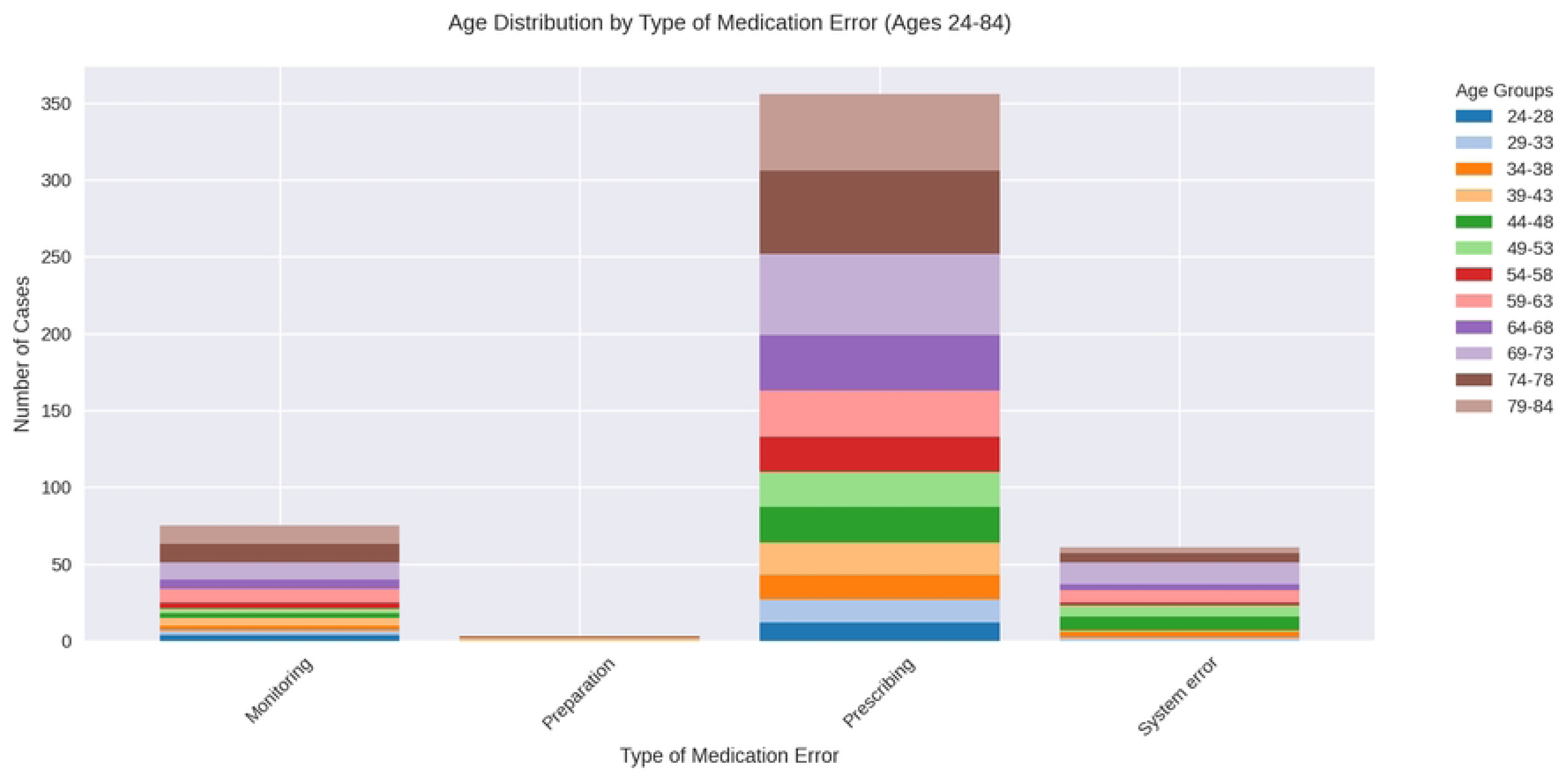
Age Distribution by Types of Medication Errors

**Table 1.** Demographic and Clinical Characteristics of Patients Experiencing Anticoagulant-Related Medication Errors.

| Variable | Category | Datix System<br>Count<br>(Percentage) | Intervention System<br>Count (Percentage) | Total<br>Count<br>(Percentage) |
| --- | --- | --- | --- | --- |
| Gender | Female | 135 (56.96%) | 197 (50.38%) | 332 (52.87%) |
|  | Male | 102 (43.04%) | 193 (49.36%) | 295 (46.97%) |
|  | Missing | 2 (0.84%) | 0 (0.00%) | 2 (0.32%) |
| Indication* | ACS | 1 (0.43%) | 2 (0.51%) | 3 (0.48%) |
|  | AF | 85 (36.17%) | 84 (21.48%) | 169 (27.00%) |
|  | APLS | 1 (0.43%) | 0 (0.00%) | 1 (0.16%) |
|  | AVR | 8 (3.40%) | 2 (0.51%) | 10 (1.60%) |
|  | CVA | 5 (2.13%) | 0 (0.00%) | 5 (0.80%) |
|  | DVT | 34 (14.47%) | 43 (11.00%) | 77 (12.30%) |
|  | HCM | 1 (0.43%) | 0 (0.00%) | 1 (0.16%) |
|  | HD | 1 (0.43%) | 0 (0.00%) | 1 (0.16%) |
|  | HRTI | 0 (0.00%) | 1 (0.26%) | 1 (0.16%) |
|  | IHD | 3 (1.28%) | 7 (1.79%) | 10 (1.60%) |
|  | LV thrombus | 6 (2.55%) | 5 (1.28%) | 11 (1.76%) |
|  | MI | 0 (0.00%) | 1 (0.26%) | 1 (0.16%) |
|  | MVR | 22 (9.36%) | 8 (2.05%) | 30 (4.79%) |
|  | PAD | 0 (0.00%) | 1 (0.26%) | 1 (0.16%) |
|  | PE | 18 (7.66%) | 40 (10.23%) | 58 (9.27%) |
|  | PVD | 0 (0.00%) | 1 (0.26%) | 1 (0.16%) |
|  | PVT | 3 (1.28%) | 0 (0.00%) | 3 (0.48%) |
|  | SMV thrombosis | 1 (0.43%) | 0 (0.00%) | 1 (0.16%) |
|  | VT prophylaxis | 45 (19.15%) | 196 (50.13%) | 241 (38.50%) |
|  | Wrong drug | 1 (0.43%) | 0 (0.00%) | 1 (0.16%) |
|  | Missing | 4 (1.70%) | 0 (0.00%) | 4 (0.64%) |
| Anticoagulant | Apixaban | 75 (31.78%) | 83 (21.23%) | 158 (25.20%) |
|  | Enoxaparin | 54 (22.88%) | 267 (68.29%) | 321 (51.20%) |
|  | Rivaroxaban | 18 (7.63%) | 16 (4.09%) | 34 (5.42%) |
|  | Warfarin | 89 (37.71%) | 25 (6.39%) | 114 (18.18%) |
|  | Missing | 3 (1.27%) | 0 (0.00%) | 3 (0.48%) |
| Type of medication error | Dispensing | 1 (0.42%) | 0 (0.00%) | 1 (0.16%) |
|  | Monitoring | 6 (2.54%) | 91 (23.27%) | 97 (15.47%) |
|  | Preparation | 3 (1.27%) | 0 (0.00%) | 3 (0.48%) |
|  | Prescribing | 158 (66.95%) | 295 (75.45%) | 453 (72.25%) |
|  | System error | 68 (28.81%) | 5 (1.28%) | 73 (11.64%) |
|  | Missing | 3 (1.27%) | 0 (0.00%) | 3 (0.48%) |
| Sub-type of error | Drug-drug interaction | 4 (1.72%) | 9 (2.31%) | 13 (2.09%) |
|  | Duplicate therapy | 43 (18.53%) | 18 (4.62%) | 61 (9.81%) |
|  | Expiry medication | 1 (0.43%) | 0 (0.00%) | 1 (0.16%) |
|  | Incorrect dose | 129 (55.60%) | 252 (64.62%) | 381 (61.25%) |
|  | Incorrect drug | 14 (6.03%) | 14 (3.59%) | 28 (4.50%) |
|  | Incorrect duration | 9 (3.88%) | 12 (3.08%) | 21 (3.38%) |
|  | Incorrect frequency | 27 (11.64%) | 25 (6.41%) | 52 (8.36%) |
|  | Incorrect quantity | 5 (2.16%) | 1 (0.26%) | 6 (0.96%) |
|  | Omission error | 0 (0.00%) | 59 (15.13%) | 59 (9.49%) |
|  | Missing | 7 (3.02%) | 1 (0.26%) | 8 (1.29%) |
| Severity of medication error<sup>+</sup> | A | 4 (1.70%) | 0 (0.00%) | 4 (0.64%) |
|  | B | 189 (80.43%) | 383 (97.95%) | 572 (91.37%) |
|  | C | 42 (17.87%) | 8 (2.05%) | 50 (7.99%) |
|  | Missing | 4 (1.70%) | 0 (0.00%) | 4 (0.64%) |
| Contributing factor | Communication issue | 15 (6.33%) | 20 (5.12%) | 35 (5.57%) |
|  | Lack of knowledge | 80 (33.76%) | 169 (43.22%) | 249 (39.65%) |
|  | Monitoring failures | 30 (12.66%) | 180 (46.04%) | 210 (33.44%) |
|  | Use of technology(CDSS) | 112 (47.26%) | 22 (5.63%) | 134 (21.34%) |
|  | Missing | 2 (0.84%) | 0 (0.00%) | 2 (0.32%) |

\*ACS: Acute Coronary Syndrome; AF: Atrial Fibrillation; APLS: Antiphospholipid Syndrome; AVR:
Aortic Valve Replacement; CVA: Cerebrovascular Accident; DVT: Deep Vein Thrombosis; HCM:
Hypertrophic Cardiomyopathy; HD: Hemodialysis; HRTI: High Risk Traumatic Injury; IHD: Ischemic
Heart Disease; LV thrombus: Left Ventricular Thrombus; MI: Myocardial Infarction; MVR: Mitral
Valve Replacement; PAD: Peripheral Arterial Disease; PE: Pulmonary Embolism; PVD: Peripheral
Vascular Disease; PVT: Portal Vein Thrombosis; SMV thrombosis: Superior Mesenteric Vein
Thrombosis; VT prophylaxis: Venous Thrombosis Prophylaxis
<sup>+</sup> A: Circumstances or events that have the capacity to cause error; B: An error occurred but the error
did not reach the patient; C: An error occurred that reached the patient but did not cause patient harm

The most common indication for anticoagulation therapy was venous thrombosis prophylaxis (38.5%, n = 241), followed by atrial fibrillation (27.00%, n = 169), and deep vein thrombosis (12.30%, n = 77), as detailed in Table 1. Notably, the distribution of indications varied between the two reporting systems, with atrial fibrillation being more frequently reported in the Datix system (36.17% vs. 21.48%), while venous thrombosis prophylaxis was more commonly reported in the Intervention system (50.13% vs. 19.15%).

### Types and Subtypes of Medication Errors

The distribution of medication errors across different anticoagulants and error types is shown in Table 2. Prescribing errors were the most prevalent (n = 453, 72.4%), followed by monitoring errors (n = 97, 15.5%) and system errors (n = 73, 11.7%). As presented in Table 2, enoxaparin was associated with the highest number of errors (n = 321, 51.2%), followed by apixaban (n = 158, 25.2%) and warfarin (n = 114, 18.2%).

**Table 2.** Distribution of Medication Errors by Anticoagulant and Error Type.

|  | Type of Medication Error |
| --- | --- |

| Anticoagulant | Monitoring | Prescribing | System error | Dispensing | Preparation | TOTAL |
| --- | --- | --- | --- | --- | --- | --- |
| **Enoxaparin** | 75 (23.4%) | 245<br>(76.3%) | 1 (0.3%) | 0 (0.0%) | 0 (0.0%) | 321<br>(51.2%) |
| **Apixaban** | 17 (10.8%) | 138<br>(87.3%) | 2 (1.3%) | 1 (0.6%) | 0 (0.0%) | 158<br>(25.2%) |
| **Warfarin** | 3 (2.6%) | 42 (36.8%) | 68 (59.6%) | 0 (0.0%) | 1 (0.9%) | 114<br>(18.2%) |
| **Rivaroxaban** | 2 (6.1%) | 28 (84.8%) | 2 (6.1%) | 0 (0.0%) | 1 (3.0%) | 33 (5.4%) |
| **TOTAL** | 97 (15.5%) | 453<br>(72.4%) | 73 (11.7%) | 1 (0.2%) | 2 (0.3%) | 626 |

Table 3 illustrates the frequency of subtypes of medication errors. Among prescribing errors, the most common subtypes were incorrect dose (n = 238, 53%), duplicate therapy (n = 54, 12%), and omission of required medication (n = 49, 10.9%), as detailed in Table 3. For monitoring and system errors, the most common subtype was also incorrect dose (n = 74, 77.1%; n = 69, 94.5%, respectively). Further analysis of incorrect dosing errors revealed that 58.7% (n = 145) involved underdosing, 32.8% (n = 81) involved overdosing, and 8.5% (n = 21) involved inappropriate dosing frequency.

**Table 3.** Error Subtypes Within the Medication Errors.

| Medication<br>Error<br>Type | Error Subtype | Frequency<br>(%) |
| --- | --- | --- |
| Dispensing | Duplicate therapy | 1 (100%) |
| Monitoring | Duplicate therapy | 4 (4.2%) |
|  | Incorrect dose | 74 (77.1%) |
|  | Incorrect duration | 2 (2.1%) |
|  | Incorrect frequency | 7 (7.3%) |
|  | Omission error | 9 (9.4%) |
| Preparation | Expiry medication | 1 (33.3%) |
|  | Incorrect drug | 2 (66.7%) |
| Prescribing | Drug-drug interaction | 13 (2.9%) |
|  | Duplicate therapy | 54 (12%) |
|  | Incorrect dose | 238 (53%) |
|  | Incorrect drug | 25 (5.6%) |
|  | Incorrect duration | 19 (4.2%) |
|  | Incorrect frequency | 45 (10%) |
|  | Incorrect quantity | 6 (1.3%) |
|  | Omission error | 49 (10.9%) |
| System error | Duplicate therapy | 2 (2.7%) |
|  | Incorrect dose | 69 (94.5%) |
|  | Incorrect drug | 1 (1.4%) |
|  | Omission error | 1 (1.4%) |

### Severity of Medication Errors

The severity of medication errors was classified according to the National Coordinating Council for Medication Error Reporting and Prevention (NCC MERP) index. The distribution of error severity is presented in Table 4 and visually depicted in Fig 2, which shows the relationship between patient age and error severity. As shown in Table 4, the majority of medication errors were classified as category B (n = 571, 91.4%), followed by category C (n = 50, 8%) and category A (n = 4, 0.6%).

**Fig 2.**
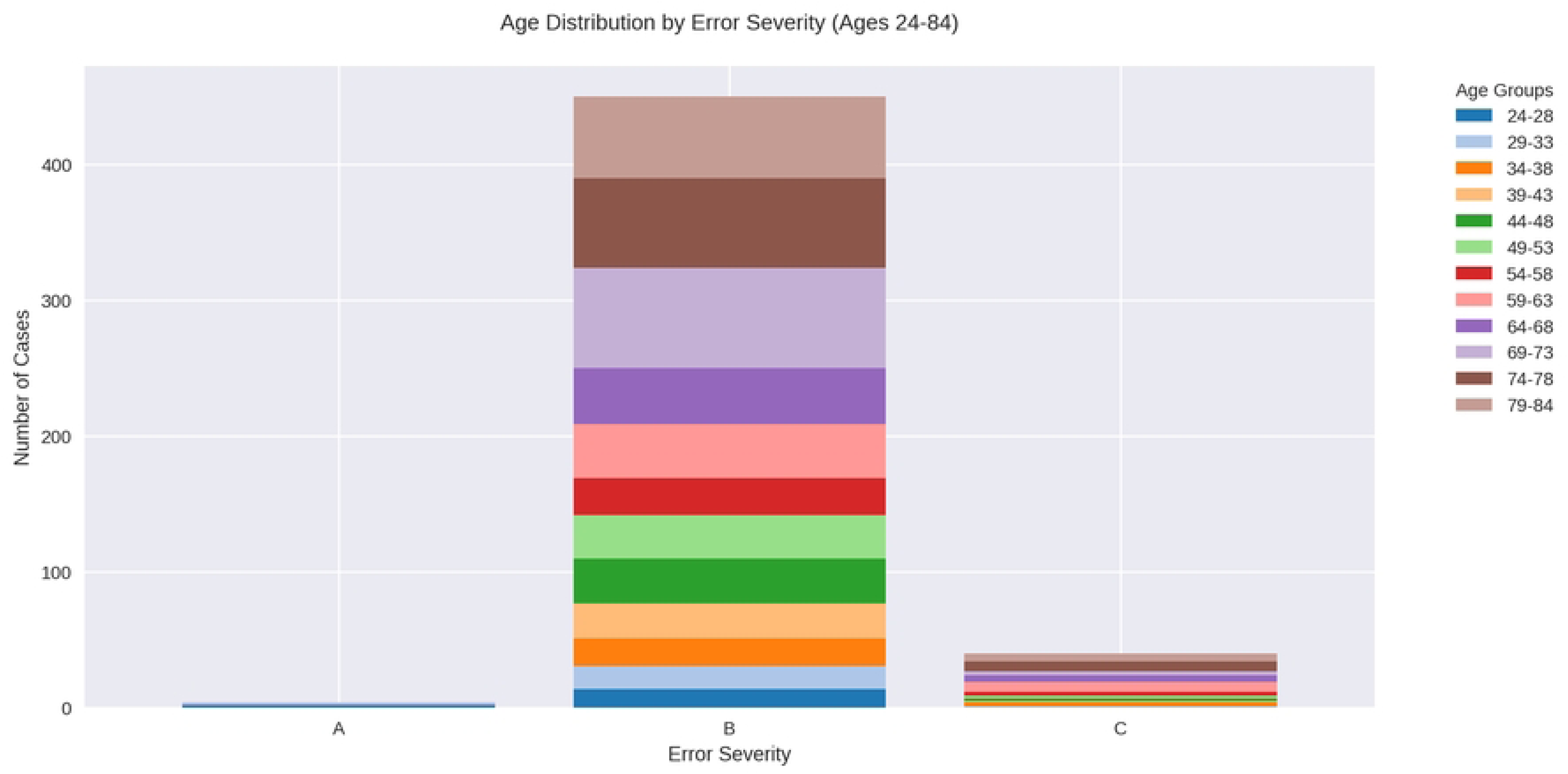
Age Distribution by Error Severity

**Table 4.**
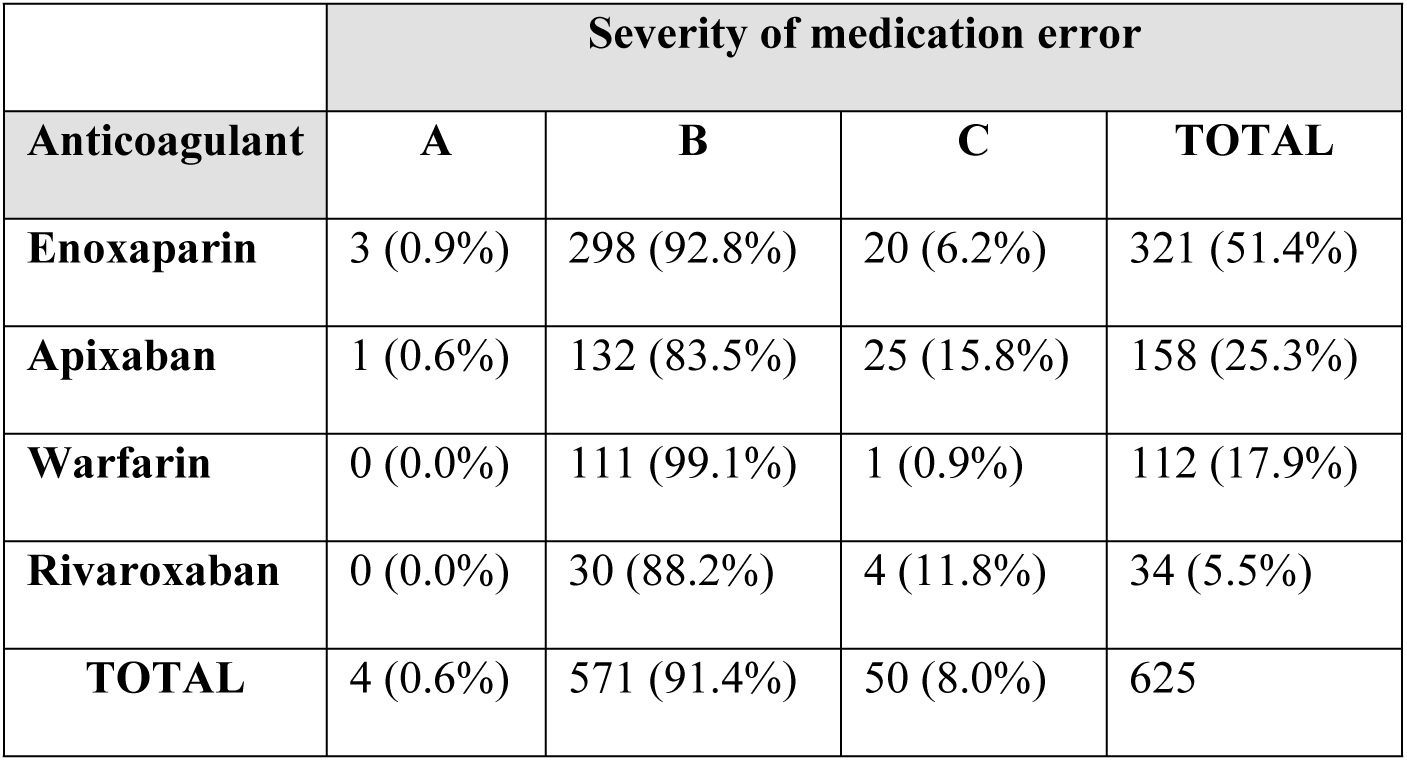
Severity of Medication Errors by Anticoagulant Type.

Enoxaparin was associated with the highest number of category B errors (n = 298, 92.8%), whereas apixaban had the highest proportion of category C errors (n = 25, 15.8%), as detailed in Table 4. As shown in Fig 2, there is a notable relationship between patient age and error severity, with older patients more frequently experiencing category B and C errors (p < 0.001).

### Relationship Between Patient Age and Medication Error Characteristics

Age distribution analysis revealed significant associations between patient age and both the severity and type of medication errors. As depicted in Fig 1, the overall age distribution shows a concentration of errors among elderly patients, with the highest frequency occurring between 70-84 years of age (n = 217, 34.4% of all errors). Patients who experienced prescribing errors tended to be older (mean age 72.1 ± 13.6) compared to those who experienced other types of errors (p < 0.001). This age-related pattern was consistent across all anticoagulant types, suggesting that age is an independent risk factor for specific types of medication errors.

Additionally, as illustrated in Fig 2, the mean age of patients experiencing category A errors (33.6 ± 8.2 years) was significantly lower than those experiencing category B (68.3 ± 14.7 years) or C errors (71.2 ± 15.5 years) (p < 0.001). Logistic regression analysis confirmed that for each decade increase in age, the odds of experiencing a major error increased by 1.42 (95% CI: 1.28-1.57, p < 0.001) after adjusting for gender, comorbidities, and anticoagulant type.

### Contributing Factors to Medication Errors and Anticoagulants

Table 5 presents the identified contributing factors to medication errors involving anticoagulants. Lack of knowledge was the predominant contributing factor, accounting for 39.7% (248/625) of all cases, followed by monitoring failure at 33.4% (209/625), as shown in Table 5.

**Table 5.** Contributing Factors to Anticoagulants and Types of Medication Errors.

|  |  | Contributing Factor |  |  |  |  |
| --- | --- | --- | --- | --- | --- | --- |
|  |  | Monitoring failure | Communication | Lack of knowledge | Use of technology (CDSS) | TOTAL |
| Anticoagulant | Apixaban | 37 (23.6%) | 12 (7.6%) | 82 (52.2%) | 26 (16.6%) | 157<br>(25.1%) |
|  | Enoxaparin | 158 (49.2%) | 13 (4.0%) | 134 (41.7%) | 16 (5.0%) | 321<br>(51.4%) |
|  | Rivaroxaban | 3 (8.8%) | 1 (2.9%) | 16 (47.1%) | 14 (41.2%) | 34 (5.4%) |
|  | Warfarin | 11 (9.7%) | 8 (7.1%) | 16 (14.2%) | 78 (69.0%) | 113<br>(18.1%) |
|  | TOTAL | 209 (33.4%) | 34 (5.4%) | 248 (39.7%) | 134 (21.4%) | 625<br>(100.0%) |
| Medication Error Type | Dispensing | 0 (0.0%) | 0 (0.0%) | 0 (0.0%) | 1 (100.0%) | 1 (0.2%) |
|  | Monitoring | 94 (96.9%) | 1 (1.0%) | 2 (2.1%) | 0 (0.0%) | 97<br>(15.5%) |
|  | Preparation | 1 (33.3%) | 1 (33.3%) | 1 (33.3%) | 0 (0.0%) | 3 (0.5%) |
|  | Prescribing | 114 (25.3%) | 33 (7.3%) | 245 (54.3%) | 59 (13.1%) | 451<br>(72.2%) |
|  | System error | 0 (0.0%) | 0 (0.0%) | 0 (0.0%) | 73 (100.0%) | 73<br>(11.7%) |
|  | TOTAL | 209 (33.4%) | 35 (5.6%) | 248 (39.7%) | 133 (21.3%) | 625<br>(100.0%) |

Chi-square analysis revealed significant associations between contributing factors and both medication error types (χ² = 504.21, df = 12, p < 0.001) and anticoagulant medications (χ² = 250.53, df = 12, p < 0.001). In the analysis of error types, as detailed in Table 5, lack of knowledge was predominantly associated with prescribing errors (245/451, 54.3%). Monitoring failure (209/625, 33.4%) was strongly linked to monitoring errors (94/97, 96.9%). Use of technology (133/625, 21.3%) showed a distinct pattern, accounting for all system errors (73/73, 100%).

Lack of knowledge was also the leading contributing factor across most anticoagulants, particularly with apixaban (52.2%, 82/157) and enoxaparin (41.7%, 134/321), as presented in Table 5. However, the use of technology factor was predominantly associated with warfarin errors (69.0%, 78/113). Monitoring failure issues showed varying patterns, being more frequent with enoxaparin (49.2%, 158/321) compared to other anticoagulants.

## Discussion

This retrospective cohort study examined anticoagulant-related medication errors in a tertiary care hospital in Saudi Arabia, identifying patterns, contributing factors, and associations with patient characteristics. Our findings revealed that prescribing errors were the most common type of medication error, with incorrect dosing being the predominant subtype. Enoxaparin was associated with the highest number of errors, and lack of knowledge emerged as the leading contributing factor. Elderly patients were particularly vulnerable to anticoagulant-related medication errors, with a significant association observed between advanced age and error severity.

### Types and Patterns of Medication Errors

Our study found that prescribing errors constituted 72.4% of all anticoagulant-related medication errors, followed by monitoring errors (15.5%) and system errors (11.7%). This predominance of prescribing errors aligns with findings from several international studies. Dreijer et al. reported that prescribing errors accounted for 68.3% of anticoagulant-related medication errors across multiple hospitals in the Netherlands [20]. Similarly, a systematic review by Alrowily et al. found that prescribing errors represented 59.7% of all DOAC-related medication errors [21]. The high prevalence of prescribing errors may reflect the complexity of anticoagulant dosing regimens, which often require adjustments based on patient-specific factors such as age, weight, renal function, and concomitant medications.

Within the category of prescribing errors, incorrect dosing was the most common subtype (53%), consistent with findings from previous studies. Henriksen et al. analyzed data from the Danish Patient Safety Database and found that 55.8% of anticoagulant-related medication errors involved incorrect dosing [22]. Similarly, a multicenter study by Schillig et al. reported that dosing errors accounted for 49.7% of all anticoagulant-related medication errors [23]. The high frequency of dosing errors may be attributed to the narrow therapeutic index of anticoagulants, complex dosing protocols, and the need for dose adjustments based on multiple patient factors.

Our analysis of incorrect dosing errors revealed that underdosing (58.7%) was more common than overdosing (32.8%). This finding differs from some previous studies, such as that by Piazza et al., which reported a higher prevalence of overdosing errors (59.4%) compared to underdosing (40.6%) [24]. This discrepancy may reflect differences in prescribing practices, patient populations, or institutional protocols. The predominance of underdosing in our study could indicate a cautious approach by prescribers, potentially due to concerns about bleeding risks, particularly in elderly patients who constituted a significant proportion of our study population.

### Anticoagulant Types and Associated Errors

Enoxaparin was associated with the highest number of medication errors (51.2%), followed by apixaban (25.2%) and warfarin (18.2%). The high error rate with enoxaparin may be attributed to its widespread use for venous thromboembolism prophylaxis, which was the most common indication for anticoagulation in our study population. Similar findings were reported by Fanikos et al., who found that low molecular weight heparins accounted for 48.3% of anticoagulant-related medication errors in their institution [25]. The complexity of weight-based dosing for enoxaparin and the need for dose adjustments in renal impairment may contribute to the high error rate.

Interestingly, while warfarin has traditionally been associated with a high risk of medication errors due to its narrow therapeutic index and numerous drug-drug interactions, it accounted for a smaller proportion of errors in our study compared to enoxaparin and apixaban. This finding may reflect the declining use of warfarin in favor of DOACs, as well as the implementation of specialized anticoagulation management services for warfarin patients in many institutions. Schillig et al. similarly reported a decreasing trend in warfarin-related medication errors following the introduction of DOACs [23].

The pattern of errors varied across different anticoagulants. Prescribing errors were predominant for enoxaparin (76.3%) and apixaban (87.3%), while system errors were more common for warfarin (59.6%). The high prevalence of system errors with warfarin may be related to challenges in integrating complex monitoring requirements and dose adjustments into electronic health record systems. Schiff et al. highlighted similar challenges with computerized physician order entry systems for warfarin, noting that these systems often lack sophisticated decision support for anticoagulant management [26].

### Error Severity and Patient Outcomes

The majority of medication errors in our study were classified as category B (91.4%) according to the NCC MERP index, indicating that the error reached the patient but did not cause harm. This finding is consistent with a study by Desai et al., which reported that 89.3% of anticoagulant-related medication errors did not result in patient harm [27]. However, it is important to note that even non-harmful errors represent system failures that could potentially lead to adverse outcomes in different circumstances.

Category C errors, which reached the patient and required monitoring to confirm that no harm occurred, accounted for 8% of all errors in our study. Apixaban had the highest proportion of category C errors (15.8%), suggesting that errors involving this medication may be more likely to require additional monitoring. This finding aligns with a study by Barr and Epps, which identified a higher risk of clinically significant errors with DOACs compared to warfarin, possibly due to less frequent monitoring and fewer opportunities to detect errors before they reach the patient [28].

### Age-Related Patterns and Risk Factors

Our study revealed a significant association between patient age and medication error characteristics. Elderly patients (70-84 years) experienced the highest frequency of errors, and patients who experienced prescribing errors tended to be older (mean age 72.1 ± 13.6 years) compared to those who experienced other types of errors. This age-related pattern is consistent with findings from previous studies. Desai et al. reported that patients aged 75 years and older had a 1.9-fold increased risk of experiencing anticoagulant-related medication errors compared to younger patients [27]. Similarly, a systematic review by Donaldson et al. identified advanced age as a significant risk factor for medication errors across various therapeutic categories, including anticoagulants [29].

The increased vulnerability of elderly patients to medication errors may be attributed to several factors, including polypharmacy, multiple comorbidities, altered pharmacokinetics and pharmacodynamics, and cognitive impairment. Older patients often require more complex medication regimens, increasing the potential for errors. Additionally, age-related changes in renal function may necessitate dose adjustments for anticoagulants, adding another layer of complexity to prescribing decisions. Our logistic regression analysis confirmed that for each decade increase in age, the odds of experiencing a major error increased by 1.42 (95% CI: 1.28-1.57) after adjusting for gender, comorbidities, and anticoagulant type. This finding underscores the importance of targeted interventions to reduce medication errors in elderly patients receiving anticoagulant therapy.

### Contributing Factors to Medication Errors

Lack of knowledge emerged as the predominant contributing factor to anticoagulant-related medication errors in our study, accounting for 39.7% of all cases. This finding is consistent with a systematic review by Salmasi et al., which identified knowledge deficits as a major contributor to medication errors across various healthcare settings [30]. The complexity of anticoagulant therapy, including dosing regimens, monitoring requirements, and drug interactions, may contribute to knowledge gaps among healthcare providers.

Monitoring failure was the second most common contributing factor (33.4%), particularly for enoxaparin errors (49.2%). This finding highlights the importance of systematic approaches to monitoring patients on anticoagulant therapy, including regular assessment of renal function, coagulation parameters, and clinical response. Fanikos et al. similarly identified inadequate monitoring as a significant contributor to anticoagulant-related adverse events, emphasizing the need for standardized monitoring protocols [25].

The use of technology, particularly clinical decision support systems (CDSS), was identified as a contributing factor in 21.3% of errors, predominantly associated with warfarin (69.0%). While technology is often implemented to reduce medication errors, poorly designed systems or alert fatigue may paradoxically increase error risk. Schiff et al. analyzed computerized physician order entry-related medication errors and found that system design flaws, including inappropriate default settings and insufficient decision support, contributed to anticoagulant-related errors [26]. This finding underscores the importance of thoughtful implementation and continuous evaluation of technology-based interventions to ensure they enhance rather than hinder medication safety.

### Implications for Practice

Our findings have several important implications for clinical practice and medication safety initiatives. First, the high prevalence of prescribing errors, particularly incorrect dosing, highlights the need for enhanced education and training for healthcare providers on anticoagulant management. This could include targeted educational programs, clinical decision support tools, and standardized protocols for anticoagulant prescribing and monitoring.

Second, the significant association between advanced age and medication errors suggests that special attention should be given to elderly patients receiving anticoagulant therapy. Comprehensive geriatric assessments, medication reconciliation, and close monitoring may help reduce error risk in this vulnerable population.

Third, the varying patterns of errors across different anticoagulants indicate that medication safety initiatives should be tailored to the specific challenges associated with each agent. For example, strategies to reduce enoxaparin-related errors might focus on weight-based dosing and renal dose adjustments, while approaches to warfarin safety might emphasize improved integration of monitoring data into electronic health record systems.

Finally, the identification of lack of knowledge and monitoring failure as key contributing factors suggests that multifaceted interventions addressing both provider education and system-level monitoring processes may be most effective in reducing anticoagulant-related medication errors.

### Limitations

This study has several limitations that should be acknowledged. First, as a retrospective study relying on reported medication errors, it may underestimate the true incidence of errors due to underreporting. Healthcare providers may be reluctant to report errors, particularly those that do not result in patient harm, leading to potential selection bias. Second, the study was conducted at a single tertiary care hospital in Saudi Arabia, which may limit the generalizability of findings to other healthcare settings or regions with different prescribing practices, patient populations, or healthcare systems. Third, while we identified associations between various factors and medication errors, the retrospective design precludes the establishment of causal relationships. Prospective studies would be needed to confirm these associations and evaluate the effectiveness of targeted interventions. Fourth, our analysis focused primarily on the characteristics and contributing factors of medication errors, with limited data on patient outcomes beyond the NCC MERP severity classification. Future studies should include more detailed assessments of clinical outcomes, including bleeding events, thromboembolic complications, and healthcare utilization.

## Conclusions

This comprehensive analysis of anticoagulant-related medication errors in a tertiary care hospital in Saudi Arabia highlights the predominance of prescribing errors, particularly incorrect dosing, and identifies lack of knowledge and monitoring failure as key contributing factors. The findings underscore the vulnerability of elderly patients to medication errors and reveal distinct patterns of errors across different anticoagulant agents. These insights can inform the development of targeted interventions to enhance medication safety for patients receiving anticoagulant therapy, ultimately improving clinical outcomes and reducing healthcare costs associated with preventable adverse events.

## Data Availability

All relevant data are within the manuscript and its Supporting Information files.

NA

## Acknowledgments

None.

